# Stereotactic arrhythmia radioablation (STAR) in patients with ventricular tachycardia: a meta-analysis of efficacy and safety outcomes

**DOI:** 10.1101/2020.05.09.20094763

**Authors:** Gustavo Arruda Viani, Juliana Fernandes Pavoni, Ligia Issa De Fendi

**Author notes:** **Correspondent author information** Gustavo Arruda Viani, Dr. Rubem Aloysio Moreira, number 155, Zip code-14021686.

## Abstract

**Objectives:** The effectiveness and safety of STAR in patients with refractory ventricular tachycardia (VT) to catheter ablation are limited to small series. We performed a meta-analysis of observational studies to summarize existing data about efficacy and toxicity following START for VT. Methods: Eligible studies were identified on Medline, Embase, the Cochrane Library, and the proceedings of annual meetings through March 2020. We followed the PRISMA and MOOSE guidelines. An estimative of % VT burden reduction at 6 months higher than 85% was considered effective. A rate of any grade 3 or higher toxicity lower than 10% and no grade 4 or 5 were considered safe. Results: Four observational studies with a total of 39 patients treated were included. The % of VT burden reduction at 6 months was 91% (CI95% 83 - 10%). The consumption of lower than 2 anti-arrhythmia drugs (AAD) at 6 months was 81%. The ejection fraction improved in 12.8%, unchanged 82%, and decreased by 5.2%. The overall survival (OS) was 92% and 82 % in 6 and 12 months. The cardiac death and disease-specific survival at 12 months were 12% and 88.5%. Late grade 3 toxicity 5% with no grade 4-5. Conclusion: STAR produced satisfactory % of VT burden reduction, with a significant reduction in the consumption of AAD at 6 months, and no severe toxicity. These findings support the continued work to develop new trials and to adopt STAR as a treatment option for medical practice.

## Introduction

Stereotactic body radiotherapy (SBRT) employs the system of 3-dimensional localization of a target to deliver an extremely high dose of radiation to a precise site even with the target in movement(1). The high precision with the system allows delivering a high dose to a specific target with no significant damage to the healthy tissue surrounding the target(1). Therefore, the SBRT has a high capability to be used for ablating tissue noninvasively, and, recently, the SBRT has been assessed for the treatment of malignant arrhythmias, mainly, for ventricular tachycardia (VT)(2,3).

VT is a potentially life-threatening arrhythmia due to a reentry circuit within a ventricular substrate, which may conduct into ventricular fibrillation, cardiac arrest, and death(4). Coronary artery disease or other non-ischemic dilated cardiomyopathy (DCM) are the leading causes related to the development of VT(4).

The treatment of VT involves antiarrhythmic drugs (AAD), and interventional therapies. The ADD has limited efficacy and several side effects, which limits its use (5). The catheter ablation with radiofrequency (RF) energy is the most frequent intervention used int the clinical practice to the cessation of the VT (6). It heats the tissue in direct contact or around to the tip of a catheter electrode, provoking tissue necrosis, which may conduct to disruption of the underlying arrhythmia substrate (4,5).

The ablation success is multifactorial and depends on the procedure types and if the underlying substrate is located deeply in the myocardium surface (7). In recent years, due to inadequate ability to disrupt some of these deep substrates has motivated the development of alternative ablative modalities such as SBRT (8).

Stereotactic arrhythmia radioablation (STAR) is a novel noninvasive SBRT application performed in an outpatient environment with no anesthesia (2,3,9). STAR is precisely direct to the VT target identified by electroanatomic mapping combined with several images and delivers high-dose radiation (usually 25- 30 Gy) to the VT target tissue point causing local tissue destruction resulting in ischemia (10,11). The initial results of some case series demonstrate a powerful antiarrhythmic effect to control the VT storm, allowing the reduction of antiarrhythmic drugs (9,12-16). However, a small study recently published showed a disappointing outcome with STAR after twelve months of follow-up for VT cessation (17).

Therefore, we designed a meta-analysis of case series and prospective studies available in the medical literature to appraise the efficacy and safety of STAR for VT, to support the continued research in the field.

## Material and methods

We conducted this systematic review and meta-analysis according to the Preferred Reporting Items for Systematic Reviews and Meta-analysis (PRISMA) statement and the Meta-analyses Of Observational Studies in Epidemiology (MOOSE) guideline(18,19). Two reviewers performed the research, selected the articles initially by title and abstract, and then read the full articles.

A systematic search was conducted by two of the investigators in PubMed, the Cochrane Central Register of Controlled Trials, and Embase for studies assessing the treatment outcomes of STAR for patients with VT from any cause. The search terms are described in supplementary table-1.

## Study selection

We included only published studies evaluating the treatment outcomes of STAR with the SBRT technique for patients with VT. Case series (5 or more patients), retrospective, or prospective studies reporting their outcome with six months of follow up or longer were included.

## Patients

We included studies of patients with VT from the ischemic, non-ischemic cardiac disease who were submitted to STAR to the cessation of VT. Studies including patients ≥18 years of age, with ≥3 episodes VT using ≥1 AAD and ≥1 catheter ablation or with contraindication to catheter ablation, were included.

## Intervention

We evaluated the efficacy and safety of STAR to the cessation of VT using any radioablation dose. Any type of technique (intensity-modulated radiotherapy or VMAT) or any type of RT machine (Cyberknife or LINAC) was allowed. Any kind of cardiac imaging to identify regions of anatomical scarring such as; single-photon emission CT (SPECT) or contrast-enhanced cardiac MRI combined or not with electroanatomical mapping information to build a VT target to radioablate were allowed.

## Outcomes

Primary outcomes were % of reduction of VT burden at six months, % of AAD < 2 at six months, and overall survival (OS) at 6 and 12 months. Secondary outcomes were the number of implantable cardio-verter defibrillator (ICD) shocks at 6 and 12 months, cardiac death, and disease specific survival rate at 12 months. The number of VT episodes pre and post-procedure was used to calculate the % reduction of VT. The calculation was performed using the same period before and after the procedure from data collected by ICD. The rate of patients consuming < 2 the AAD at 6 months was calculated by the proportion of patients taking none or one AAD at 6 months. The cardiac ejection fraction (EF) was considered improved if after the procedure a sustained increased >5% from the baseline. The EF was unchanged if after the procedure the EF continued in the baseline. The EF was decreased when a sustained drop of 5% below the baseline was detected. The toxicity analysis was performed considering grade 3 or higher toxicity as a severe complication from STAR to cessation VT. Any toxicity grade 3 or higher related or probable to the STAR was considered as an event. Any reporting of ICD malfunction was also registered.

## Clinical data

The data from the patient, treatment characteristics, and outcomes for all studies included were retrieved. Two reviewers independently selected data using a standardized method. The following information was collected: author, year, study design, number of patients, STAR dose, RT machine (LINAC vs. Cyberknife), % New York class III/IV, ejection fraction (EF) pre-procedure, % of patients using > 1 AAD, any toxicity grade 3 or higher, treatment time, PTV volume, follow-up time and clinical outcomes.

## Data synthesis and analysis

The rates of events of each outcome were calculated using the proportion rate (PR, i.e., %VT burden reduction) of patients who developed the outcome of interest with the 95% confidence interval, and the 12 statistic assessed statistical heterogeneity. An 12 value of lower than 25% was interpreted as a low level of heterogeneity. We used the random-effect model due to a relevant variation in studies’ characteristics. The % of VT burden reduction at 6 months of 70% was considered ineffective for the STAR in this meta-analysis, whereas a % of VT burden reduction at 6 months of 85% or higher was considered satisfactory. We considered as safe for the STAR if any toxicity grade 3 or higher was lower than < 10%, and no grade 4-5. Based on a binomial distribution with a one-sided type I error of 0.05, type II error of 0.10 (power of at least 90%), a sample size higher than 36 patients would be necessary pooling the studies to validate the efficacy and safety of STAR. A p-value lower than 0.05 was considered significant in all analyses. The meta-analysis was performed using the Open Meta-Analyst free open software. The quality of case series was evaluated by a tool developed by Murad et al. that consist of eight items that can be categorized into four domains: selection (1 question: 1 point), ascertainment (two questions: 2 points), causality (four questions: 4 points), and reporting (1 question: 1 point)(20).

## Results

We identified in our searches 236 studies reporting the outcomes of STAR to VT refractory after catheter ablation. After applying the inclusion criteria, we selected 3 case series studies and 1 prospective cohort (phase 1/II study), including 39 patients with VT refractory who were treated with STAR reporting the outcomes with a median follow-up of 14 months (10-28 months)(17,21-23). Supplementary figure 1 describes the search strategy and the reasons for the exclusion of some studies.

All studies were published from 2017 to 2020. The sample size ranged from 5 to 19, the median age was 66 (63 - 66) years, with a median of pre-ejection fraction ranging from 23 to 26.5%, and with the majority of included patients with ischemic cardiac disease 70%(40-80%), table-1 summarized the clinical characteristics of patients of studies. Two studies performed STAR with a Cyberknife and two with LINAC, employing the same dose 25 Gy/1 fraction. Two studies combined electroanatomic mapping with imaging (CT, MRI, and PET) and CT planning to draw the target, and the other two used electroanatomical mapping with CT planning. All studies used an ITV based on the motion of respiratory cycles from 4D CT or breath-holding CT, table-2 provide technical aspects of studies. All studies used similar workflows to perform the STAR as described in figure-1.

**Figure-1.**
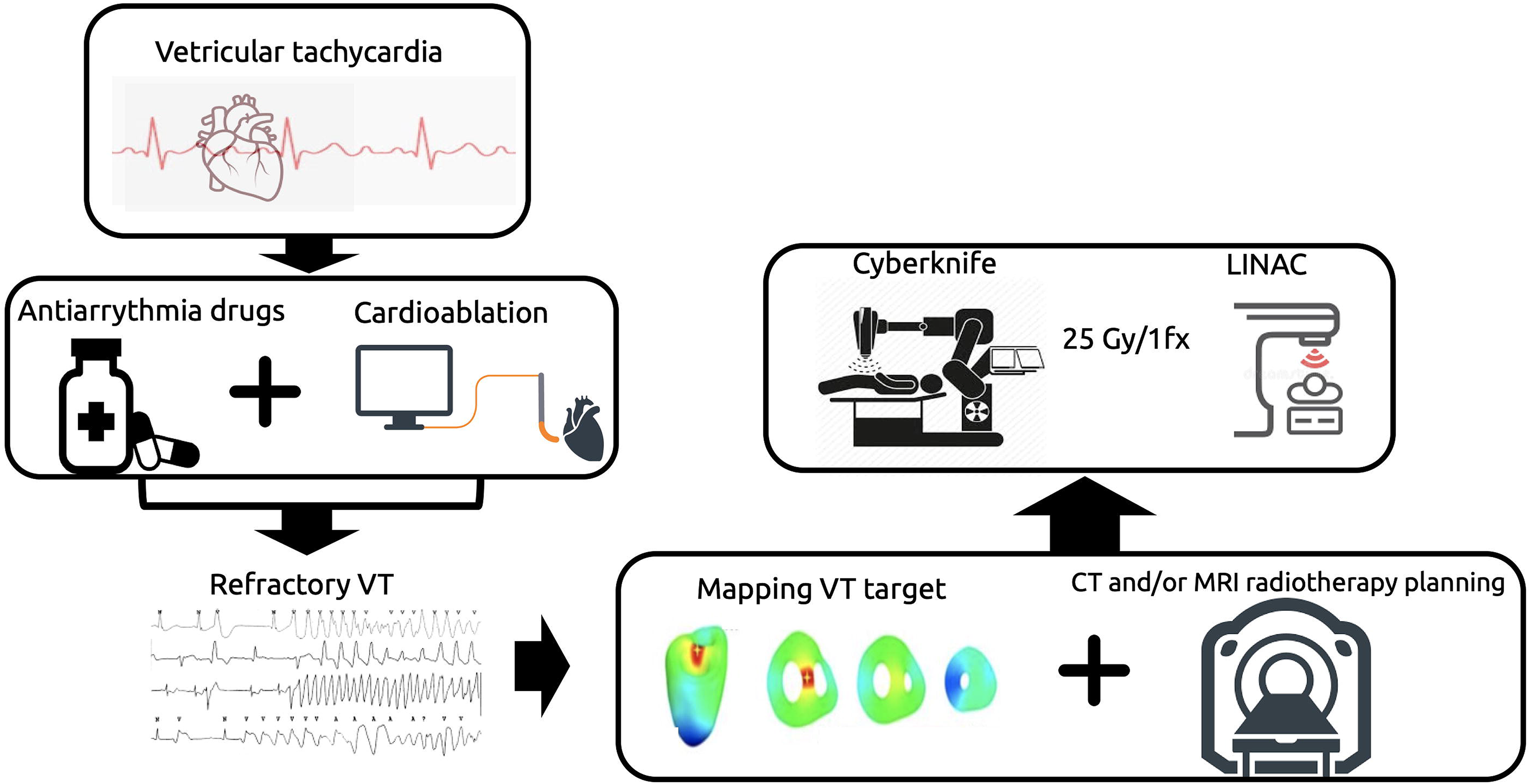
Summarized description of the Workflow of patients to perform STAR: 1-patients selection: inclusion of patients with VT refractory to catheter ablation and using AAD; 2-electroanatomic mapping (3-dimensional electroanatomical or electrocardiogram (ECG) imaging map); 4-Anatomic scar imaging (MRI, CT, Nuclear imaging); 5-Creation of a CTV identifying arrhythmogenic scar substrate using electroanatomical mapping and anatomical scar imaging; 6-Study of the motion with 4D CT or with CT breath-holding to create the ITV(LINAC); 7-Imaging fiducial marker for respiratory tracking (existing ICD lead or insertion of a temporary) for CyberKnife, and creation of an ITV with respiratory/cardiac motion; 8. Development of a radiation treatment plan targeting scar 9. Positioning, simulation with imaging, and aligning of the patient 10. Treatment with 25 Gy/1 fraction on the targeted area from a LINAC or Cyberknife.

## % VT burden reduction, % AAD and OS at 6 months

Four studies reported the primary endpoint as an outcome. Pooling the % VT burden reduction resulted in a reduction of 91% (CI95% 83 - 100%) at 6 months, figure-2a, with no heterogeneity. The % consuming < 2 of AAD at 6 months pooling the three studies was 80% (CI95% 50-100), figure 2b. The OS rate at 6 and 12 months was 92% (CI95% 83-100) and 82% (CI95% 65-98) respectively, as demonstrated in figure 2c and 2d.

**Figure-2.**
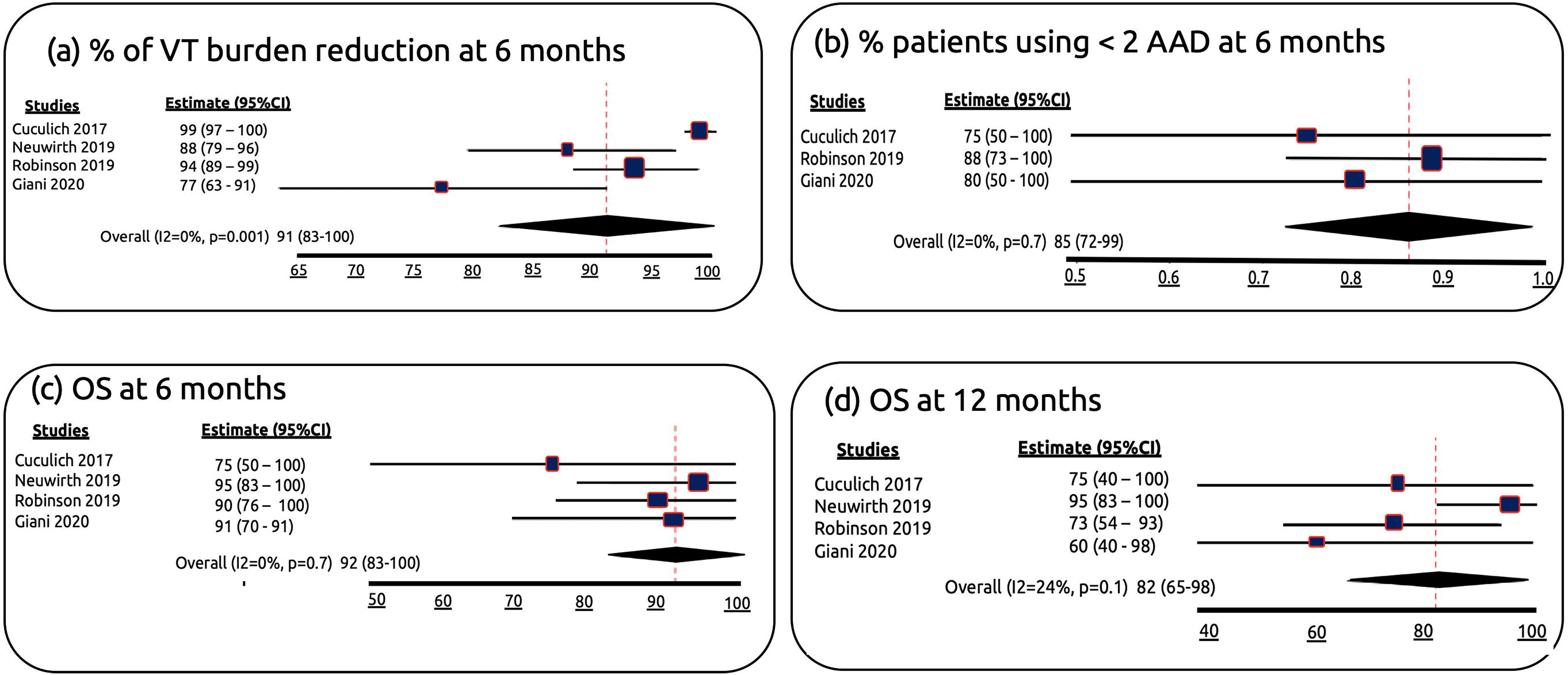
(a) % VT burden reduction at 6 months (b) consuming < 2 of AAD at 6 months (c) OS at 6 months (d) OS at 12 months.

## ICD shock, Cardiac death, and cardiac specific survival

Three studies reported the ICD shock as an outcome. Pooling the absolute number of ICD shock pre and post STAR at 6 months, a reduction of 90% was verified (100 vs 10, p=0.001), figure 3a, and comparing ICD shock at 12 months after STAR the pooling data for the reduction was 28% (100 vs 72, p=0.03), figure-3b. The cardiac death rate and cardiac specific survival at 12 months were 12.5% and 88.5% respectively, as demonstrated in figure 4a.

**Figure-3.**
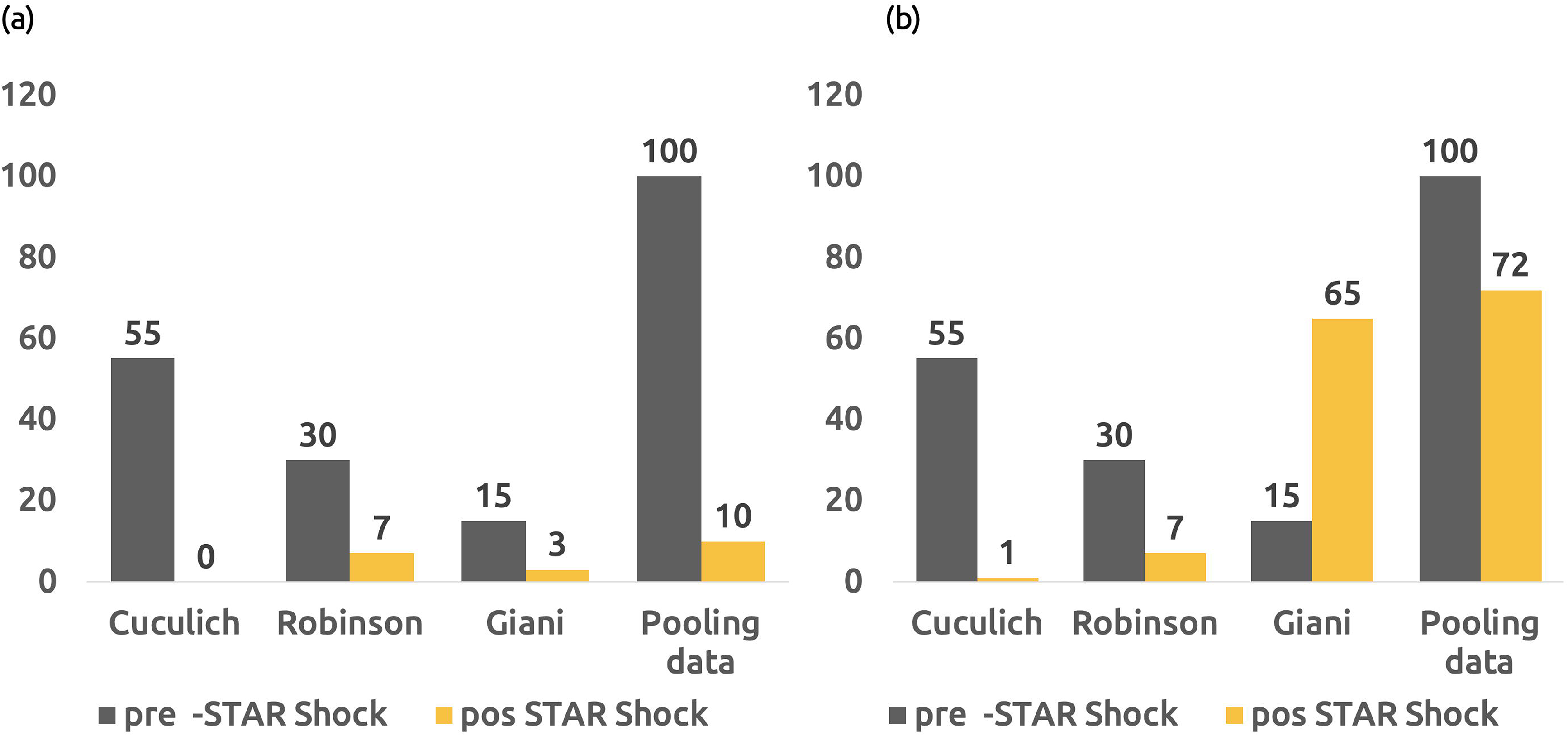
(a) Absolut number of shock event at 6 months after STAR. (b) Absolut number of shock event at 12 months after STAR

## Cardiac Ejection fraction after STAR

Four studies reported the ejection fraction before the treatment. Pooling the data, 12.8%, 82%, and 5.2% had EF improved, EF unchanged and EF decreased, respectively, as demonstrated in figure 4b. Two patients from Gianni study were considered as EF decreased. Both patients’ failure from the radioablation with posterior VT storm reducing the EF, i.e., the worst in the EF was not directly related to the STAR(17).

**Figure-4.**
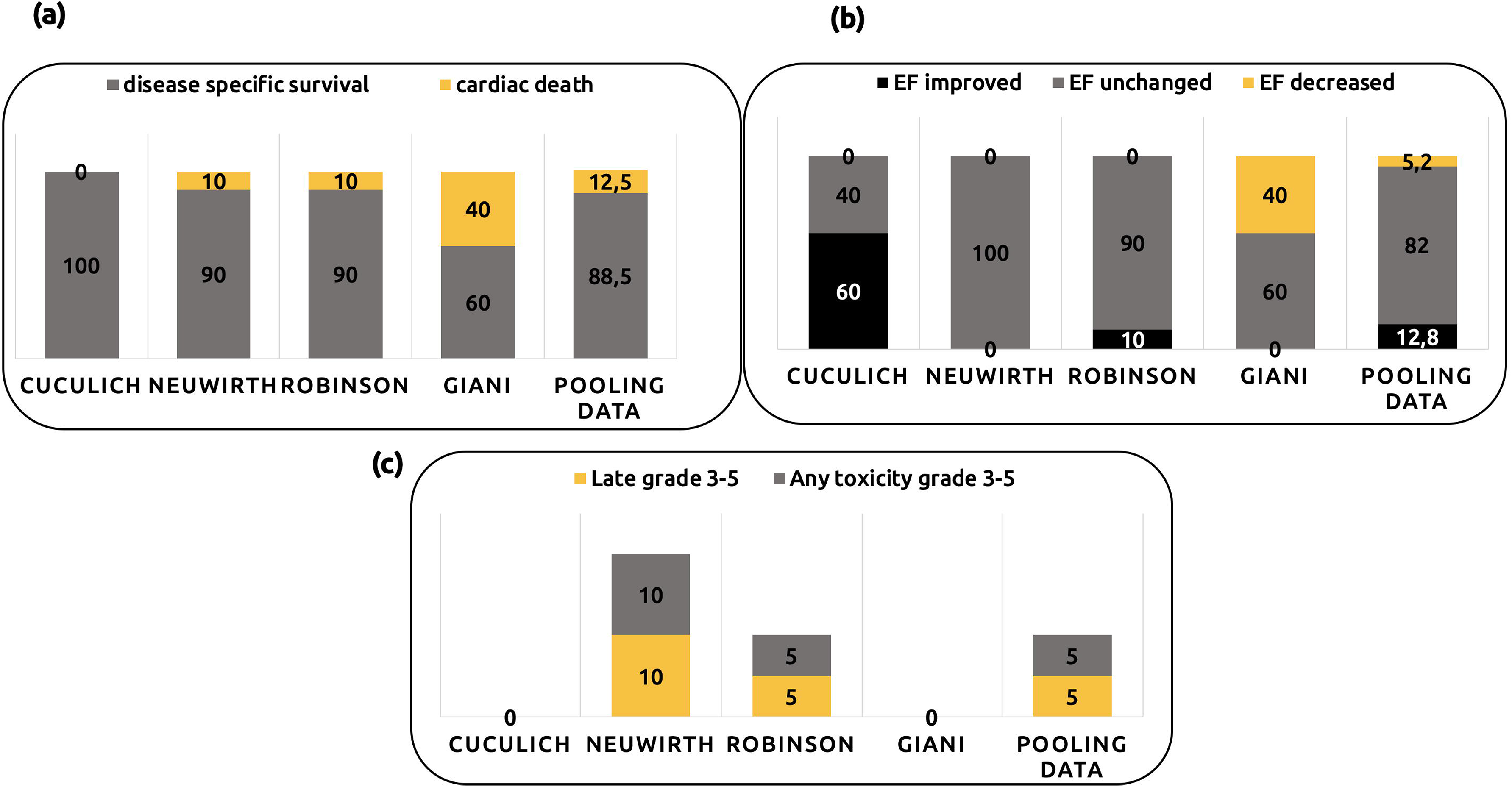
(a) % VT burden reduction at 6 months (b) consuming <2 of AAD at 6 months (c) OS at 6 months (d) OS at 12 months.

## Toxicity and ICD functioning

All studies reported the toxicity and ICD status post procedure. No acute grade 3-5 toxicity was reported in all studies. Late grade 3-5 was reported in two patients, both with grade 3 toxicity. One patient was considered grade 3 late toxicity due to worst of mitral regurgitation after 17months after radiosurgery in the Neuwirth et al. study(23). Another patient had a pericardial effusion documented as possibly related grade 3 in the Robison et al. study(21). No grade 4-5 toxicity related to the procedure was reported. Pooling the data, the rate of grade 3 toxicity was 5%, as shown in figure 4c. No reporting of ICD malfunctioning was reported after the STAR.

## Quality of case series

Using the tool created by Murad et al.(20) and giving 1 point for each question, all case series achieved 8 points, with an agreement of 100% between the reviewers, being considered of high-quality reports.

## Discussion

Case series are studies with uncontrolled design and recognized risk of bias(24,25). However, this kind of study has profoundly influenced the medical literature to continue developing our knowledge(25). Meta-analysis is a sort of study centered on a research problem that tries to identify, assess, and synthesize all the research evidence available. However, a lot of medical areas exhibit an absence of a high level of evidence, such as a randomized controlled trial(26). Consequently, in clinical scenarios where a new medical procedure is developed or situations involving rare disease, case series are the only source of evidence to guide decisions and future researches(27). Recently, the meta-analysis of case series has been considered as a valuable tool to handle clinical problems like that(26). The recent development of using SBRT to treat patients with VT refractory to catheter cardio ablation is at this point since the majority of initial evidence comes from case reports or case series(9,12-17,21-23,28,29). Hence, we designed a meta-analysis to assess the efficacy and safety of STAR, including all the case series describing the efficacy and toxicity related to the procedure.

The present study is the first meta-analysis to include case studies of STAR for VT in patients refractory to catheter ablation. We excluded the case reports of our analysis to reduce the risk of publication bias. In total, we excluded 8 case reports published between 2015-2020, including a total of 8 patients treated by STAR(9,12-16,28,29). The decision in limiting the meta-analysis to the case series had a direct impact on the number of studies collected. So, we decide to estimate a minimal sample size required to give reliability to the outcomes using data from the literature as limits. The % VT burden reduction at 6 months, combining the four studies was 91% (CI95% 83%-100%), which was superior to the 85% previously specified. All studies used VT burden collected by ICD as a primary endpoint; thus, we decide to utilize it as a primary endpoint to estimate the STAR efficacy. Three studies adopted the reduction/discontinuation of AAD as a criterion for effectiveness. Pooling the rate of consuming at 6 months, we observed 85% of patients consuming < 2 the AAD after STAR. In the study leading by Neuwirth et al., other endpoints related to time-event were also assessed, such as time to shock, time to the antiarrhythmic storm, and time to anti-tachycardia pacing (23). All these endpoints were considered significant and favorable for STAR, but, unfortunately, we cannot compare it with other studies. The definition of success is fundamental to evaluate new procedures. The VT burden reduction is an important endpoint, and currently, can be considered as the marker of success, but their relationship with time can also provide valuable information about the effect durability from the procedure for in the future to be compared with other ablation procedures (radiofrequency ablation, cryoablation). Although the inclusion criteria have some variation among the studies (supplementary table-2), the patients’ characteristics were similar with reduced variability, which translates to a similar OS rate at 6 months. Of note, Gianni et al. included patients with reduced expected survival, and the OS at 12 months was significantly distinct from the other studies. In the same study, the VT burden reduction at 6 months was favorable; however, after 12 months, the rate had a drop down to 40%, with two patients dying from cardiac failure, being considered a disappointing outcome(17). The possible explanations for this disappointing outcome pass through the inclusion of very high-risk patients, technical difficulties to target delineation with suboptimal coverage, and dislocation of ICD to guide the treatment. From our data, the rate of cardiac death at 12 months from Gianni et al. study was 4 times higher than others (figure-3a). Our data agree with the experience of researches from Emory University using STAR for critically ill cardiac patients(30). We excluded the Emory case series of our analysis because of the short follow-up (<6 months). The study included 10 patients, all of them refractory to catheter ablation (median 2 previous ablation), taking > = 2 AAD, besides that, 3 patients had left ventricular assist devices and 1 patient with intra-aortic balloon pump. In their analysis, STAR reduced the total ICD shocks in 68% (2.9 shocks/month pretreatment and 0.9 shocks/month posttreatment). These data were lower than of our pooling data (90% at 6 months) but are comparable to the Gianni study with 40% at 12 months and call attention to a better selection of patients who benefit from STAR, and the need of longer follow-up. All the studies used a similar workflow to perform the procedure, as summarized in figure-1. The SBRT technique to cardioablation must consider the respiratory motion, while precisely track the VT target throughout both the cardiac/respiratory cycle.

The source of ventricular arrhythmia often occurs in areas of myocardial scar(2,4). In general, during the invasive cardiac ablations, areas of the scar can be recognized using electroanatomic mapping systems. In these mappings, the scars are identified as low-voltage areas. In addition to electroanatomic mapping, several imaging techniques have been used to identify myocardial scars such as CT, echocardiography, nuclear perfusion imaging, PET, or contrast-enhanced MRI(21,22). All information provided by the electroanatomic mapping and images is useful for accurately drawing the targets in CT planning for SBRT. These technical aspects were similar among the studies to perform the STAR, as described in table-2 and figure-1. At this point, we cannot establish differences among the studies to recommend the most accurate combination of mapping and imaging to draw the target. However, further research in this field, combining the target location with both cardiac and respiratory cycle, could provide additional information to avoid under dose during the intrafraction period in such regions giving additional margins to accommodate the PTV adequately.

The late side effects are concerned, mainly due to the high radiation dose delivered and the high risk of other complications as the cardiac dysfunction with valve stricture, pericardiac effusion, papillary muscle dysfunction or new conduction abnormalities in such fragile population. Considering these risks, we pre-specified a grade 3 acute/late toxicity lower than 10% and no grade 4-5 for the STAR be considered safe. The outcomes for acute/late toxicity with 5% of grade 3 and no grade 4-5 toxicity observed in the studies can be considered safe. Another point of concern is the malfunction of ICD during or after the STAR. We did not find any report on the STAR leading malfunction of the ICD, which also confirm the safety of the procedure.

Further studies employing real-time target localization with magnetic resonance imaging-guided is a promising technology with an enormous capability to improve the therapeutic index for VT ablation(29).

## Conclusion

Our meta-analysis shows that STAR is effective and safe to treat selected patients with VT refractory to catheter ablation and AAD. The level of reduction of VT burden was superior to the prespecified level of 85%, with no significant differences between Cyberknife and LINAC. The STAR also was effective in reducing the consumption of AAD and the number of ICD shock. Although the studies have had a similar criterion of inclusion, the selection of patients is crucial to guarantee acceptable outcomes, once studies including patients more fragile or critically ill, the rate of success dropdown. The workflow employed to perform STAR among the studies was clear and similar, demonstrating a high homogeneity considering STAR a procedure with a short life. The main challenge of STAR is the accurate definition and delineation of the target, and in this systematic review, we were incapable of detecting differences between studies that combined several images with electroanatomical mapping or only used electroanatomical mapping. Nevertheless, given the complex motion of both cardiac and respiratory cycle, the precise definition of the target is extremely critical that the radiation oncologist and electrophysiologist use the maximum information available to guarantee or improve the therapeutic success of the procedure. Further researches are needed to understand the relationship between the target, cardiac/respiratory movement, and margin to create the ITV to avoid an under coverage the substrate. STAR with a dose 25 Gy/1fx was the standard and resulted in a grade 3 acute / late toxicity low (5%) with no grade 4-5, which confirm its safety. It is crucial to highlight that no reporting of ICD malfunctioning was registered using a machine with 6MV or 10 MV during or after the procedure. Finally, the findings showed in this meta-analysis provide a solid base for the continued conduction of prospective studies employing STAR in this clinical scenario.

## Data Availability

All data referred to in our manuscript are collected from published studies and all of them are available on the electronic database.

## Figure and table legends

Table-1 Characteristic of patients included in the studies.

Table-2 Technical aspects of radiotherapy used in the studies for STAR of VT.

## Supplementary material

Figure-1 Flowchart of studies included.

Table-1 Search terms employed to find the studies.

Table-2 Inclusion criteria of studies using STAR for VT.

## Notes

### Competing Interest Statement

The authors have declared no competing interest.

### Funding Statement

No funding

